# Obstructive sleep apnea, positive airway pressure treatment, and postoperative delirium: protocol for a retrospective observational study

**DOI:** 10.1101/19002501

**Authors:** Christopher R King, Krisztina Escallier, Yo-El S. Ju, Nan Lin, Ben Julian A. Palanca, Sherry McKinnon, Michael S Avidan

## Abstract

**Introduction:** Obstructive sleep apnea (OSA) is common among older surgical patients, and delirium is a frequent and serious postoperative complication. Emerging evidence suggests that OSA increases the risk for postoperative delirium. We hypothesize that OSA is an independent risk factor for postoperative delirium, and that in patients with OSA, perioperative adherence to positive airway pressure (PAP) therapy decreases the incidence of postoperative delirium and its sequelae. The proposed retrospective cohort analysis study will use existing datasets to: (i) describe and compare the incidence of postoperative delirium in surgical patients based on OSA diagnosis and treatment with PAP; (ii) assess whether preoperatively untreated OSA is independently associated with postoperative delirium; and (iii) explore whether preoperatively untreated OSA is independently associated with worse postoperative quality of life. The findings of this study will inform on the potential utility and approach of an interventional trial aimed at preventing postoperative delirium in patients with diagnosed and undiagnosed OSA.

**Methods and Analysis:** Observational data from existing electronic databases will be used, including over 100,000 surgical patients and ∼10,000 intensive care unit (ICU) admissions. We will obtain the incidence of postoperative delirium in adults admitted postoperatively to the ICU who underwent structured preoperative assessment, including OSA diagnosis and screening. We will use doubly robust propensity score methods to assess whether untreated OSA independently predicts postoperative delirium. Using similar methodology, we will assess if untreated OSA independently predicts worse postoperative quality of life.

**Ethics and dissemination:** This study has been approved by the Human Research Protection Office at Washington University School of Medicine. We will publish the results in a peer-reviewed venue. Because the data is secondary and high risk for re-identification, we will not publicly share the data. Data will be destroyed after 1 year of completion of active IRB approved projects.

**Article summary:** **Strengths and limitations of this study, (containing 5 short bullet points, no longer than one sentence each, that relate specifically to the methods)**

- Our granular database includes routine structured preoperative screening for OSA, processed laboratory results, and verified comorbid diagnoses.
- We have limited information on the severity of most comorbidities, creating the possibility for substantial residual confounding.
- Our database includes near-universal and standardized nurse-driven delirium evaluations at multiple time-points as well as clinician diagnoses.
- Compared to prior studies, the large sample size will allow for more aggressive confounder adjustment utilizing linked structured medical histories, intraoperative records, and administrative data.
- Selection bias and confounding by indication are important limitations, which we will address using advanced statistical methods.

## INTRODUCTION

Delirium is described in the Diagnostic and Statistical Manual of Mental Disorders, 5^th^ Edition as a disturbance in attention, awareness, and cognition that develops over a short period of time and tends to fluctuate in severity over the course of a day.^1^ It is a common postoperative complication with important costs. The reported incidence of postoperative delirium in older adults ranges from 10-70%, depending on context.^2^ Patients with postoperative delirium require longer intensive care unit (ICU) stays,^3^ experience greater institutionalization and death after discharge,^4^ and report decreased quality of life (QoL).^5^ As a result, postoperative delirium is associated with a substantial increase in healthcare costs.^6 7^ Delirium has been proposed as an indicator of quality of care in older adults,^8^ and will affect an increasing proportion of patients as the population ages.

The current literature contains suggestive evidence that obstructive sleep apnea (OSA) is a common^9 10^ and independent risk factor for postoperative delirium.^11-15^ In a small prospective study, Flink et al. reported that OSA is an independent predictor of postoperative delirium in older adults undergoing total knee arthroplasty with an odds ratio of 4.2.^11^ A prospective study of 92 patients undergoing cardiac surgery found that a preoperative apnea hypopnea index of 19 or higher was associated with increased risk of postoperative delirium (odds ratio, 6.4; 95% confidence interval, 2.6 to 15.4).^15^ A large observational study found that patients with undiagnosed OSA had worse postoperative outcomes than those with diagnosed OSA.^16^ An exploratory 114-patient randomized trial of preoperative positive airway pressure (PAP) found no impact of the intervention on delirium, but did find that OSA severity predicted postoperative delirium.^12^ A retrospective study^14^ and case report^13^ also offer support for the relationship between OSA and postoperative delirium. Several plausible biological explanations for this relationship exist, including hypoxia, chronic inflammation, and disruption of normal sleep architecture as mediators.^17 18^ However, the studies linking OSA and postoperative delirium have been small, and it is important to confirm or refute the association in a larger and more diverse sample.

We have previously investigated perioperative risks conferred by OSA. In the Barnes-Jewish Apnea Prevalence in Every Admission Study (BJ-APNEAS),^19^ a cohort of 14,962 elective surgery patients, we found a 12.9% (n = 1939) prevalence of previously diagnosed OSA. Depending on the screening instrument, roughly 10-40% of patients without a diagnosis were identified as high risk for OSA.^20^ We validated a new diagnosis in about 80% of tested patients screening as high risk.^21^ Therefore, the true overall prevalence of OSA was about 20-25%. Both a history of OSA and a positive OSA screen were associated with admission to the ICU postoperatively.^19^ Patients with known OSA, but not those screening high risk, had longer ICU stays. Patients screening high risk had significantly higher 1-year mortality than those with low risk scores.^19^ However, delirium was not routinely assessed at that time. Others have found that these patients are at increased risk of serious pulmonary,^22 23^ cardiac,^14 24^ and neurological^18^ postoperative complications.

The gold standard therapy for OSA, PAP, reduces hypoxic events, reduces markers of chronic inflammation, and improves sleep.^25-27^ American Society of Anesthesiologists (ASA) practice guidelines^28^ recommend the optimization of PAP therapy prior to surgery. Unfortunately, adherence to prescribed PAP therapy is low. It is estimated that 30% of patients who have been prescribed PAP never initiate therapy,^29^ and many eventually discontinue therapy or have suboptimal adherence.^30^ At our preoperative assessment clinic, approximately 50% of surgical candidates with OSA report adherence with PAP therapy. Similarly, Guralnick et al. found that only 33% of adult surgical patients with moderate or severe OSA used PAP for ≥4 hours per night.^31^

Our proposed retrospective cohort study has two co-primary hypotheses: (i) the presence of OSA (diagnosed or suggested by high-risk screen) increases the incidence of postoperative delirium and (ii) adequate treatment of OSA with PAP therapy reduces the risk of postoperative delirium. Secondary hypotheses are (i) high-risk screenings for untreated OSA in the preoperative period are independently associated with increased risk for postoperative delirium and (ii) untreated OSA in the preoperative period is independently associated with decreased postoperative QoL.

## Methods

### Data sources and Setting

The cohort will include all adults admitted postoperatively to either our general surgical or cardiothoracic ICUs (SICU, CTICU) between August 2012 to August 2018 who have any postoperative delirium assessments and a pre-anesthesia evaluation (where our primary exposure is reported). Data from electronic medical record databases at Barnes Jewish Hospital will be obtained and combined. This will include the preoperative anesthesia assessment, preoperative laboratory values, the day-of-service inpatient record with home medications reconciliation, the intraoperative anesthesia record, the inpatient record (providers’ notes, nursing assessments, laboratory values, vital signs, medication administration record), and administrative records. Although detailed socio-economic data will not be available, we will use administrative data on insurer, race, ethnicity, and link home addresses to census-level socioeconomic measures. For some of the patients, we will also use data from our ongoing SATISFY-SOS registry study, which tracks the intermediate term postoperative health and well-being of unselected surgical patients (NCT02032030).^32^

Based on typical admissions rates to our SICU (∼3,200 patients per year) and CTICU (∼1,200 patients per year), we estimate conservatively that the final dataset will include >10,000 patients. SATISFY-SOS is a prospective registry study; we estimate that about 2,500 patients will be available for this analysis, based on enrollment and survey completion rates.

### Main Outcomes and Exposures

The main outcome will be the incidence of postoperative delirium. Several years ago, our institution implemented routine delirium assessment in our ICUs and trained all ICU nurses to administer the Confusion Assessment Method for the Intensive Care Unit (CAM-ICU).^33^ Patients in the SICU and CTICU are now assessed twice daily for delirium. Scoring on the Richmond Agitation and Sedation Scale (RASS) is also assessed regularly and recorded, typically at the same time as the CAM if it is being performed. Patients will be coded as delirious if they have any positive delirium assessment during their ICU stay. Each episode will be characterized as hyperactive (RASS >0) or hypoactive (RASS ≤0).^34^ Secondary exploratory analyses will examine for differences with delirium type. Although delirium occurs outside the ICU, at our institution it is assessed is a non-systematic fashion. To avoid selectively recorded data and ascertainment biases related to the decision to perform a CAM on the wards, we will only analyze ICU assessments. Note extraction for chart diagnoses is not possible with this dataset and billing diagnoses do not specify a chronicity.

Previous OSA-related data from our preoperative assessment clinic (**Table 1** and **Figure 1**) and published literature^19 20 35^ were used to generate the estimated numbers of patients in each category in **Figure 2**. We routinely screen with the STOP-BANG (Snoring, Tiredness, Observed Apnea, High Blood Pressure, Body Mass Index > 35 kg/m^2^, Age > 50, Neck Circumference > 40 cm, Male Gender) criteria to determine OSA risk.^20^ We shall implement recent modifications of the STOP-BANG instrument (e.g., including Age, BMI and Neck Circumference as continuous rather than dichotomous variables) that have been shown to improve its predictive value and specificity.^27 36 37^ PAP adherence is patient reported and documented in the preoperative assessment. Patients will be categorized as “adherent” if they report “routine PAP use”. We will investigate if patients with in-hospital PAP use are more similar to those with good adherence in terms of outcomes and covariates. Hours of PAP use in the ICU are recorded in the EHR; however, this outcome is a mixture of treatment for obstruction and other causes of respiratory failure and is causally dependent on intraoperative factors and postoperative mental status, so we do not intend to use it as a covariate or outcome.

**Figure 1.**
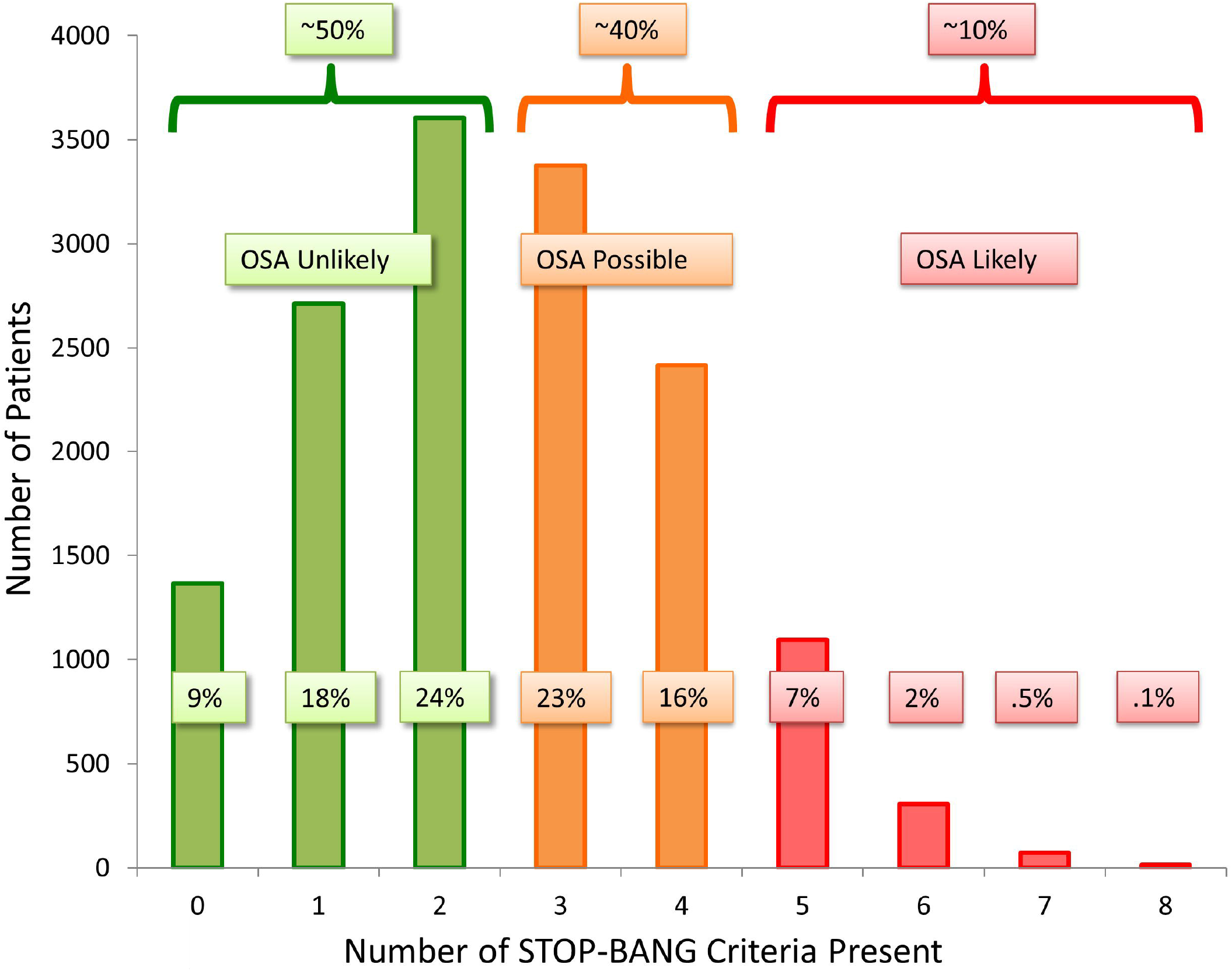
This figure shows data from 14,962 patients at our preoperative assessment clinic who did not carry a prior obstructive sleep apnea (OSA) diagnosis.^19^

**Figure 2.**
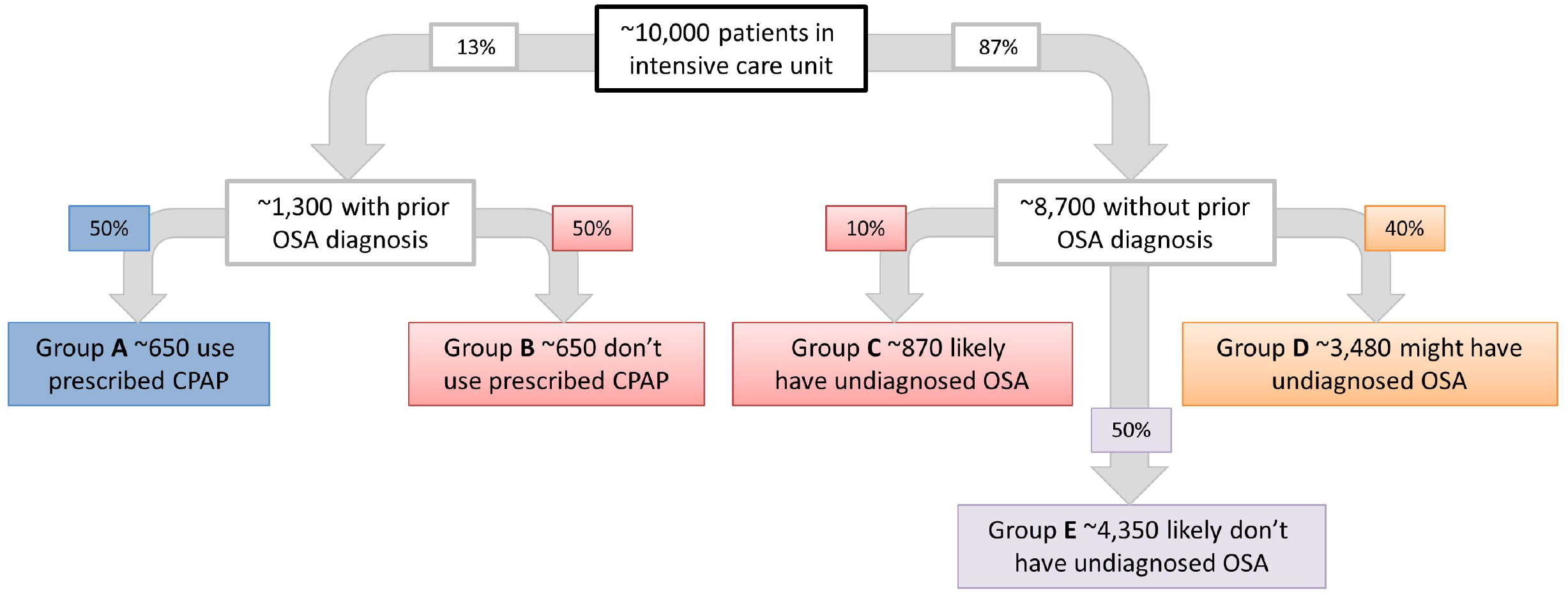
This figure shows a predicted breakdown of patients based on previous data from our preoperative assessment clinic. Approximately 1,300 (13%)^19^ of the approximately 10,000 patients in the study cohort will carry a diagnosis of obstructive sleep apnea (OSA), of whom about half (∼650) will have reported non-adherence to home PAP therapy (Group B). Of the remaining 8,700 patients, based on the current STOP-BANG criteria, about 870 (≥5 out of 8 positive criteria) are very likely to have moderate or severe undiagnosed OSA (Group C).^20^ Approximately 3,480 patients (3 or 4 positive criteria) might have undiagnosed OSA (Group D), and ∼4,350 patients (<3 positive criteria) are unlikely to have undiagnosed OSA (Group E).

| **Table 1** | **Number** | **Percent**<br>**(95% Confidence interval)** |
| --- | --- | --- |
| Adherent with treatment for obstructive sleep apnea | 477 | 51.4%<br>(48.2% to 54.6%) |
| Non-adherent with treatment for obstructive sleep apnea | 451 | 48.6%<br>(45.4% to 51.8%) |
| Out of a random sample of 7,730 patients at our preoperative assessment clinic, 1,000 carried a prior diagnosis of obstructive sleep apnea. Treatment usage was reported for 928 of these patients. Compliance was assumed only for those who reported routine usage of continuous positive airway pressure. |  |  |

SATISFY-SOS tracks intermediate-term postoperative outcomes; patients complete postoperative surveys (approximately 1 month and 1 year after surgery) that includes the Veterans Rand 12 Item Health Survey (VR-12), a validated measure of QoL. QoL will be a secondary outcome in this subset of patients; based on our prior work with SATISFY-SOS, 1500-2000 responses (versus ∼100,000 extracted the EHR) will be available for analysis.^38^ We will not link to delirium or other assessments from independent studies conducted during this period at BJH (ENGAGES, PODCAST).

### Covariates

Our models will include demographics (age, race, ethnicity, sex) as well as census-tract level economic variables. In prior work we identified several predictors of delirium: average volatile anesthetic dose, units of blood products transfused intraoperatively, and ASA physical status.^39^ EuroSCORE, a measure of severity of comorbidities, was also found to be predictive; however, it is only used for cardiac surgery, and we will substitute the Charlson comorbidity index.^40^ Other predictors will include preoperative use of sedating medications, alcohol and other intoxicants, surgery performed, baseline laboratory values (hemoglobin, creatinine, hemoglobin A1C, INR, bilirubin, albumin), baseline pain score, history of cognitive impairment, and preoperative psychiatric diagnoses. We will categorize procedures into a small number of “types” and use existing calibrations between surgery code and mortality.^41^ Based on our prior data^19^ the most common surgical types with be orthopedic (∼20%), general (∼10%), urologic (∼10%), gyecologic (∼10%), otolarygologic (∼8%), cardiothoracic (∼6%), and neurosurgical (∼6%). Our prior data do not contain good estimates of the ICU admission rates for these specialties; however, we can anticipate a substantial enrichment of cardiothoracic surgeries (at least 25%) based on the total admission rate to the CT-ICU versus SICU and a substantial decrease in neurosurgical cases as many patients are excluded from CAM measurement. Several intra- and postoperative variables will also be used: duration of surgery, duration of cardiopulmonary bypass, total intraoperative vasopresor and inotrope (norepinephrine, epinephrine, dopamine, dobutamine, phenylephrine and vasopressin) doses, intraoperative urine output, intraoperative fluids transfused, duration of coma, mechanical ventilation, use of sedatives, opioids, hypnotics, and organ dysfunction scores.^42^ SATISFY-SOS patients will additionally have multidimensional preoperative measures of anxiety, pain, functionality, stroke, visual impairment, and cognition.^43-45^

### Primary analysis plan and bias reduction

In our dataset there are no plausible sources of exogenous variation in OSA exposure or CPAP adherence to eliminate bias due to unmeasured confounders. For the primary analysis, we will use a propensity-score based approach and semi-parametric regression adjustment to reduce bias due to measured variables. We will create propensity scores for OSA diagnosis or high STOP-BANG using non-parametric regression. We will use the fitted propensity score and covariates in a flexible regression method based on an ensemble of decision trees (Bayesian Adaptive Regression Trees, BART^46^); this two-stage approach has been shown to be valid and robust,^47-49^ accounting for the uncertainty in the mechanisms of exposure and allowing nonlinear effects, interaction terms, and heterogeneity of treatment effects.^50-53^ As a sensitivity analysis we will compare the average treatment effect on the treated from our primary analysis with propensity score matching based estimates of the same with greedy 1:1 matching.^54-56^ Treatment effect estimates will be reported with 99% credible / confidence intervals. We will compare the above method to logistic regression with all variables entered linearly for the propensity and adjustment model. We will calculate a c-statistic as well as other overall fit statistics to assess the fit of this final model and will use the model to calculate odds ratios (with 99% confidence intervals) associated with each predictor. In the final regression model, statistical significance will be assumed for p values <0.01. Fitted rates in each group and the absolute risk difference (average treatment effect on linear scale) with credible interval will also be reported.

Because some variables are plausibly on the causal pathway connecting OSA and CPAP adherence and postoperative delirium (eg postoperative opioid and anxiolytic use could be less in those with untreated OSA *because they have OSA*, leading to less delirium) simply treating them as confounders would produce biased estimates ^57^ and we will initially exclude them and examine for mediation if the overall association is notable.

### Secondary Analyses

We will use a similar regression method to report variables associated with PAP adherence and in-hospital initiation of PAP. We will use a similar technique to estimate the effect of PAP on delirium given an OSA diagnosis. QoL outcomes will be handled with a similar regression model. We will also conduct exploratory analyses. For example, we will investigate possible mechanistic associations with delirium, if relevant data (e.g. oxygen saturation data) are available. We will also investigate whether outcomes are different between those who carry a diagnosis of OSA and those who screen positive for OSA. We plan to explore stratifications according to OSA severity.

### Missing Data and Loss to Follow Up

We expect that some data will be missing in the proposed study, especially as we plan to combine multiple data sources. Depending on the types, patterns and frequencies of missing variables, we will select accepted statistical approaches in order to minimize omission of patients from the analyses. Multiple imputation has been shown to be robust to the violation of normality assumptions and has produced appropriate results in similar contexts. We will conduct sensitivity analyses to evaluate the robustness of our results with and without imputation. There will be no imputation for the main risk factors of interest (OSA diagnosis or treatment) or for the primary outcome of the study (incident delirium).

For our primary outcome, loss to follow up will be a negligible problem as patients are rarely discharged while still at risk for new onset delirium. For the SATISFY-SOS cohort, efforts to minimize true loss to follow up have been described elsewhere. ^32^

### Power analysis

Based on the estimated numbers in each group in **Figure 2**, this study will be adequately powered (>80%) for the three most relevant comparisons (i.e., delirium incidence in Group A vs. Group B; Group A vs. Groups B+C; and Groups A+B+C vs. Group E). For example, for the comparison between the smallest groups (Group A vs. Group B), with one sided alpha < 0.05, there is >80% power to detect a 6% difference (from 26% observed in ENGAGES^58^ to 20%) in delirium incidence.^59^ We will not adjust the p values for multiple comparisons. However, when assessing variables for independent associations with delirium, we shall use a more stringent alpha value < 0.01.

### Ethics and Dissemination

The Human Research Protection Office at Washington University School of Medicine in St. Louis has approved this study (IRB #201311088). The conduct and reporting of this observational study will follow STROBE guidelines.^60^ Once the investigation has been completed, we intend to publish the results in a peer-reviewed publication. We also intend to present the results of this work at professional conferences for the anesthesiology community. The nature of the dataset (high resolution clinical histories linked to administrative records) makes de-identification a serious risk, and we do not plan to publicly share the data. Encryption will be used for any web-based information transmitted. The data will be stored on private protected network storage. Access will be restricted to research team members in a role-specific manner. Individual patient identifiers will be destroyed after the linking process is complete. Because the data are purely secondary, no formal data sharing is planned unless investigators obtain a separate approval for its access with Washington University’s IRB. Primary outcomes will be pre-specified, as will analytical techniques. Additional not pre-specified analyses will be treated as hypothesis-generating.

### Patient and Public Involvement

No explicit patient or public comment was sought in the design of the study. Patient-centered research has previously identified ICU delirium as a life-changing event with major consequences to quality of life; examples of patient experiences can be found at icudelirium.org. Because this is a retrospective database study, no attempt will be made to directly contact patients with the findings.

## Discussion

This large observational study will clarify if there is an independent link between OSA and postoperative delirium in the ICU. It will also show if this hypothetical increased risk is mitigated by treatment with PAP. It is important in science to replicate previous findings,^61-63^ which in the case of this study is the reported association between OSA and postoperative delirium,^11 12^ although this time in a broader surgical population. Because of its large size, this study will be useful for comparison between and among groups based on other risk factors.

This study will have important strengths compared to the existing literature, most notably the very large and granular database including routine structured preoperative screening for OSA, and postoperative delirium detection in the ICU setting. The sample size will allow for a more aggressive confounder adjustment compared to smaller studies. The population will be diverse in both comorbidities and surgery performed, allowing a more tailored identification of patients who benefit from PAP and greater generalizability. As with other large retrospective studies, purely statistical error will be small in magnitude. We have a relatively high quality assessment of medical confounders due to our experienced preoperative clinic and a well-implemented assessment of delirium reducing measurement error in key variables. We have largely pre-specified our analysis, reducing the potential for “analyst degrees of freedom” introducing spuriously high confidence after multiple comparisons. The statistical approach should provide a strong predictive model and reduce the degree of “overfitting” compared to common techniques like stepwise selection. ^64-68^

There are important limitations to the approach we are taking in this observational study. Foremost is selection bias. Patients who seek and adhere to treatment are different in many difficult to observe ways from those who do not. For example, PAP diagnosis and adherence (conditional on severity) is likely associated with socioeconomic status, care of other chronic conditions, and coping strategies. Presence of OSA or non-adherence to prescribed PAP could induce surgeons to not offer highly invasive procedure options (reducing surgical severity), cause patients to present later (increasing surgical severity), or cause patients with an otherwise lower burden of morbidity to be more aggressively admitted to the ICU where they are eligible for delirium assessments. OSA severity is likewise associated with both PAP diagnosis and adherence, making the net direction of confounding difficult to predict. Differing from selection bias, downstream indirect effects of OSA such as additional supplemental oxygen, higher usage of telemetry monitoring, and avoidance of sedating drugs may be protective. Although our preoperative clinic assessments are routinely thorough medical histories, we will have limited information on the severity of most comorbidities, leaving residual confounding. Most comorbidities are reported simultaneously, meaning that we will not be able to distinguish between confounders and mediators; simply adjusting for them may increase or decrease bias. Our intraoperative measures suffer the same difficulty.

The common problem of missing data can reduce the statistical power of a study and can produce biased estimates and invalid conclusions if severe. There are measurement errors for both the primary exposure and outcome which will decrease the validity of the associations. These analyses rely on subjective patient reporting of OSA history and PAP adherence. The STOP-BANG screening while reasonably accurate, is imperfect and may create false positives. We will try to confirm the diagnosis of OSA in our study subjects with the data available to us. Unfortunately, objective measures of PAP adherence from the actual PAP devices will not be available. Because patients tend to over-estimate their own adherence,^69 70^ we expect that using self-reported adherence will tend to under-estimate its influence on postoperative delirium rather than suggest a falsely positive association. We will attempt to obtain information from the electronic health record on in-hospital use of home PAP devices, since this may signify home adherence with PAP therapy. Treatment with alternative modalities, such as mandibular advancement devices, is not being assessed. OSA severity may be a key parameter which will be unable to obtain; others have found that apnea-hypopnea indices greater than 15 ^71^ or 30 ^72^ associated with postoperative complications. We have undertaken substantial efforts to standardize assessment of delirium in our ICUs as described above; however, there is doubtless error due to busy nursing staff and subjective elements in the assessment. Because PAP and OSA symptoms could influence delirium assessment, these errors may be informative and create additional bias.

The most rigorous way to answer whether treatment of OSA prevents postoperative delirium would be to conduct a prospective randomized, controlled trial of perioperative PAP in patients already diagnosed with OSA who are scheduled for elective surgery. However, given the established benefits of PAP in these patients, it would be unethical to randomize patients (especially those already prescribed PAP) to a non-treatment arm. Therefore, a large observational study is likely to be the most appropriate initial design for addressing this question.

Evidence of an independent risk association between untreated OSA and postoperative delirium would strongly warrant further investigation. An important question for future prospective study would be whether efforts at diagnosing OSA in the immediate preoperative period could mitigate postoperative delirium and its sequelae. We believe that this would be feasible, since we have already demonstrated within our institution that it is practical to identify patients with probable undiagnosed OSA using simple, economical screening methods.^19^ This study will further identify patients likely to benefit from focused interventions.

If we find that PAP non-adherence and untreated OSA are independent risk factors for postoperative delirium, this would inform two key priorities. First, it would reinforce the importance of promoting adherence to perioperative PAP therapy. Second, it would provide a strong impetus for conducting a randomized controlled trial in elective-surgery patients with undiagnosed OSA, which we could not ethically implement in patients who already carry a diagnosis of OSA. We hope to use the foundational work proposed in this observational study to guide the design of such a trial, with the goals of reducing postoperative delirium and improving associated outcomes for the large number of patients at risk due to OSA.

## Data Availability

Because the data is high risk for re-identification, we will not publicly share the data. Because the data are purely secondary, no formal data sharing is planned unless investigators obtain a separate approval for its access with Washington University?s IRB

## Contributors

CRK contributed to the statistical methods initial draft of protocol and critical revisions of protocol KE contributed to the overall study design and critical revisions of the protocol, YSJ, NL, BJP, SLM MSA contributed to the study design and critical revision of protocol.

## Competing interests None

### Patient consent

Not required.

### Ethics approval

Human Research Protection office, Washington University in St. Louis.

